# Philosophical Perspectives in Health Education for Hemodialysis Patients: A Scoping Review

**DOI:** 10.1101/2025.09.14.25335738

**Authors:** Atikah Fatmawati, Tintin Sukartini, Moses Glorino Rumambo Pandin

## Abstract

**Background:** Health education is crucial for hemodialysis patients, yet often limited to biomedical aspects. Philosophical perspectives, such as autonomy, caring, and humanism, may strengthen its ethical and holistic dimensions.

**Objective:** To map the integration of philosophical perspectives into health education for hemodialysis patients.

**Methods:** A scoping review was conducted using Arksey and O’Malley’s framework, refined by Levac and PRISMA-ScR guidelines. Databases searched included CINAHL (Ebsco), Scopus, Web of Science, PubMed, and ProQuest (2020–2025). Articles addressing philosophy, ethics, or humanistic theories in hemodialysis health education were included. Data were extracted and analyzed thematically.

**Results:** Four themes emerged: (1) autonomy and ethical responsibility, (2) caring ethics in nurse–patient relationships, (3) phenomenological insights into patients’ lived experiences, and (4) humanistic approaches emphasizing dignity and meaning. Few studies directly applied philosophical frameworks in structured educational interventions.

**Conclusion:** Philosophical perspectives enrich health education for hemodialysis patients by promoting autonomy, caring, and ethical engagement. Further studies are needed to develop and evaluate philosophy-informed educational models in nephrology nursing.

## Introduction

Health education is a key pillar in hemodialysis patient management, necessary to optimise adherence to therapy regimens, self-management, and quality of life. A meta-analysis found that educational interventions significantly improved adherence to diet, fluid intake, and quality of life in hemodialysis patients (Sultan & Froelicher, 2025). However, current approaches to health education still focus heavily on biomedical and technical aspects, while ethical and humanistic dimensions such as dignity, autonomy, and the therapeutic relationship are often neglected.

Research in Korea indicates that health literacy is a key factor in improving self-care behaviour in hemodialysis patients, and patient awareness of their knowledge is significantly correlated with nurses’ caring explanations (Lee & Cho, 2025). Furthermore, shared decision-making (SDM) practices are becoming increasingly important in the context of Chronic Kidney Disease (CKD), although professional training in SDM remains scarce and untested from a patient perspective (Meijers et al., 2023).

Philosophical approaches to health education open new dimensions for more reflective and person-centred nursing practice. Recent experimental research has shown that an educational program based on Jean Watson’s theory of human caring, which integrates motivational interviewing and caring values, successfully improves patient adherence to diet, fluids, and medication and reduces disease burden, with high patient satisfaction rates (Yangöz & Özer, 2025).

A study from China examined the effects of self-determination theory-based education on hemodialysis patients. Interventions that address patients’ autonomy needs, through personal interactions, scenario simulations, and dietary preference selection, have been shown to improve knowledge, self-management, and reduce anxiety and depression in hemodialysis patients (Zhang et al., 2025). This demonstrates that philosophical theories emphasising patient intrinsic autonomy make an important contribution to developing more comprehensive health education.

From a philosophical framework, the aspects of autonomy and caring can be conceptually strengthened. According to Kant’s deontological paradigm, although not widely applied directly in the context of hemodialysis, humans are viewed as rational beings with dignity, and therefore their decisions should be respected as ends, not means. Empirical evidence suggests that nurses’ autonomy contributes to enriching the moral dimension of nursing practice, similar to the egalitarian principles and ethical reflection of Tronto’s theory. Tronto, in his ethics of care, proposed five phases of caring: caring about (attention), caring for (responsibility), caregiving (concrete action), care receiving (patient response), and caring with (solidarity and trust). A recent study found that nurses’ professional autonomy influences all of these phases in the context of end-of-life care, a concept that can be transferred to the realm of hemodialysis patient education (Gilbert & Lillekroken, 2024).

However, despite the partial application of various philosophical approaches, there is little literature systematically mapping the application of philosophical concepts such as autonomy, caring, phenomenology, and humanism in hemodialysis patient health education. This philosophical foundation has the potential to enrich the ethical and existential dimensions of patient education. Therefore, this scoping review aims to map and synthesise the literature discussing the integration of philosophical perspectives in hemodialysis patient health education from 2020 to 2025.

The primary objective of this review is to provide a comprehensive overview of philosophical approaches in hemodialysis patient health education, identify dominant theories and practice models that have been used, and identify research gaps that require the development of philosophy-based interventions. Thus, this study is expected to provide theoretical and applied contributions to developing educational nursing practices that are more humanistic, ethical, and empowering for patients.

## Method

### Data Retrieval Strategy

This study is a scoping review to systematically answer the research questions and is structured according to the Preferred Reporting Items for Systematic Reviews and Meta-Analyses (PRISMA) 2020 guidelines (Page et al., 2021). This study reviews previous research on philosophical perspectives used in health education for hemodialysis patients. The detailed activities include determining data search strategies and/or information sources, selecting studies through quality assessment based on eligibility criteria and quality assessment instruments, and data synthesis and extraction.

This scoping review addresses three research questions: 1) How are philosophical perspectives (autonomy, caring, humanism, and phenomenology) integrated into health education for hemodialysis patients? 2) What philosophical theories are most commonly used? 3) What are the implications for nursing practice and patient education?

The literature search was conducted in five international databases: CINAHL (Ebsco), Scopus, Web of Science, PubMed, and ProQuest. Key words and Boolean operators used in the literature search were “nursing” OR “nurse” OR “healthcare” AND “ethics” OR “moral” OR “principles” OR “values” AND “hemodialysis” OR “dialysis” OR “renal therapy” OR “kidney treatment” AND “patient care” OR “patient rights” OR “informed consent” OR “advocacy” AND “professionalism” OR “conduct” OR “responsibility” OR “accountability”.

### Eligibility Criteria

Eligibility criteria in this study include inclusion and exclusion criteria. The inclusion criteria in this study were empirical articles, literature reviews, or conceptual studies that discuss philosophical, ethical, or humanistic frameworks in health education, quantitative, qualitative, or mixed methods studies, full-text articles, and peer-reviewed articles. Meanwhile, the exclusion criteria in this study included editorials, commentaries, and non-peer-reviewed publications that lacked philosophical discussion, as well as studies focusing solely on biomedical interventions without a connection to education or philosophy. In addition, to limit the scope of the study, the researchers used the PCC (Population, Concept, Context) method, as shown in the following table:

### Quality Assessment

Literature selection used the PRISMA (Preferred Reporting Items for Systematic Reviews and Meta-analyses) method. The PRISMA Flow Diagram in this study is shown in Figure 1.

**Figure 1.**
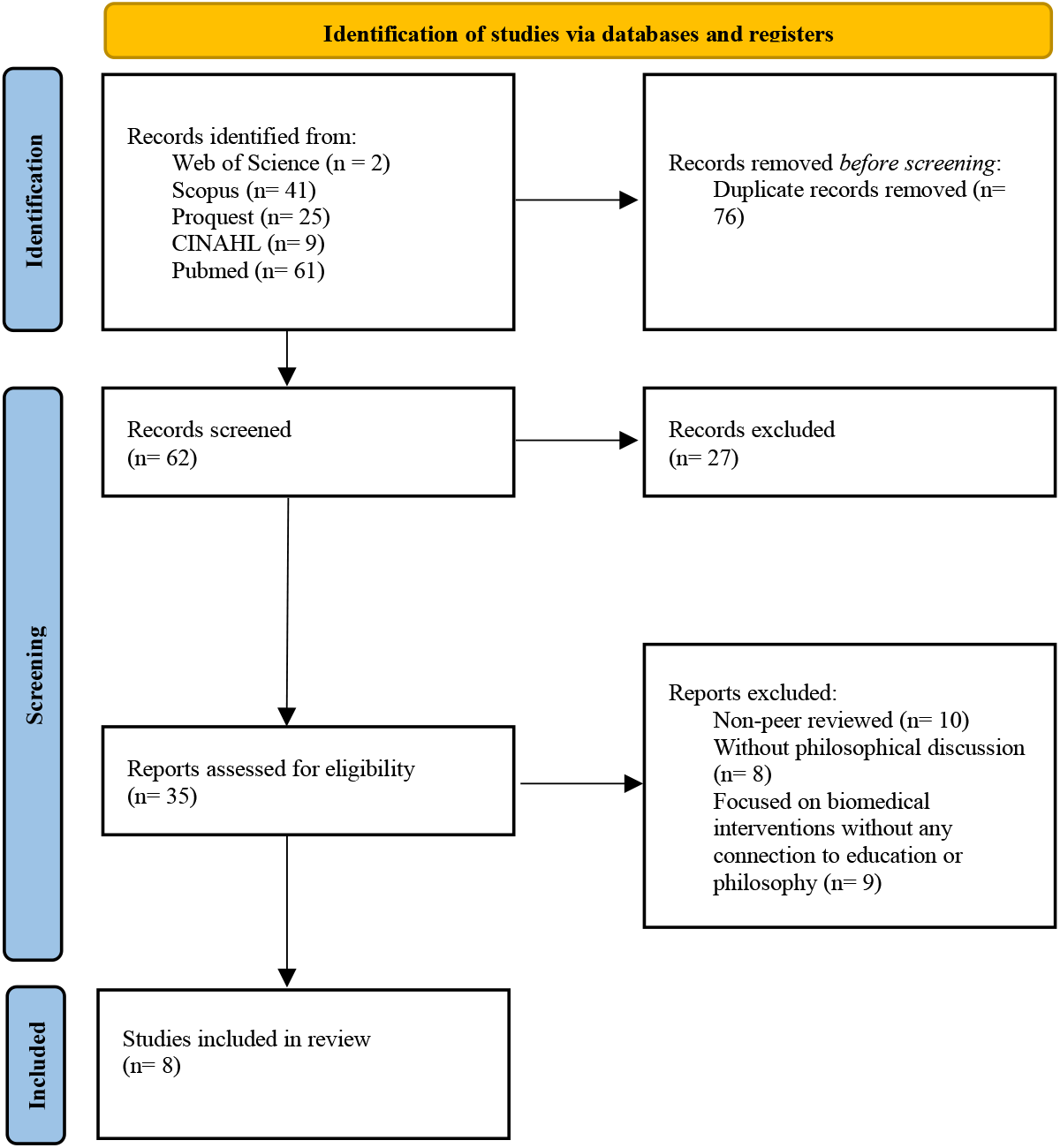
PRISMA *Flow Diagram*

### Synthesis Data

The data synthesis process in this study was conducted by comparing literature that met quality assessments and inclusion and exclusion criteria. The data synthesis refers to the research objective, which is to analyse the integration of philosophical perspectives (autonomy, caring, humanism, and phenomenology) in health education for hemodialysis patients.

### Data Extraction

The data extraction output is in the form of a table consisting of the title, researcher’s name and year of publication, study design, participants, and main findings.

## Results

Based on the selection process, the research using a scoping review technique on philosophical perspectives used in health education for hemodialysis patients identified eight articles for further analysis. A summary of the analysis of the articles on philosophical perspectives used in health education for hemodialysis patients is presented in Table 2.

**Table 1.** PCC Summary

| Componen | Keterangan |
| --- | --- |
| *Population (P)* | Hemodialysis patients (adults) undergoing regular therapy, including those with end-stage chronic kidney disease (ESRD). |
| *Concept (C)* | Philosophical perspectives in health education, including: deontological ethics (Kantian autonomy), caring ethics (Watson, Tronto), humanism, phenomenology/existentialism, and self-determination theory |
| *Context (C)* | Healthcare settings (hospitals, hemodialysis clinics, or community settings) with a focus on health education, nursing interventions, or patient education programs. |

**Table 2.**
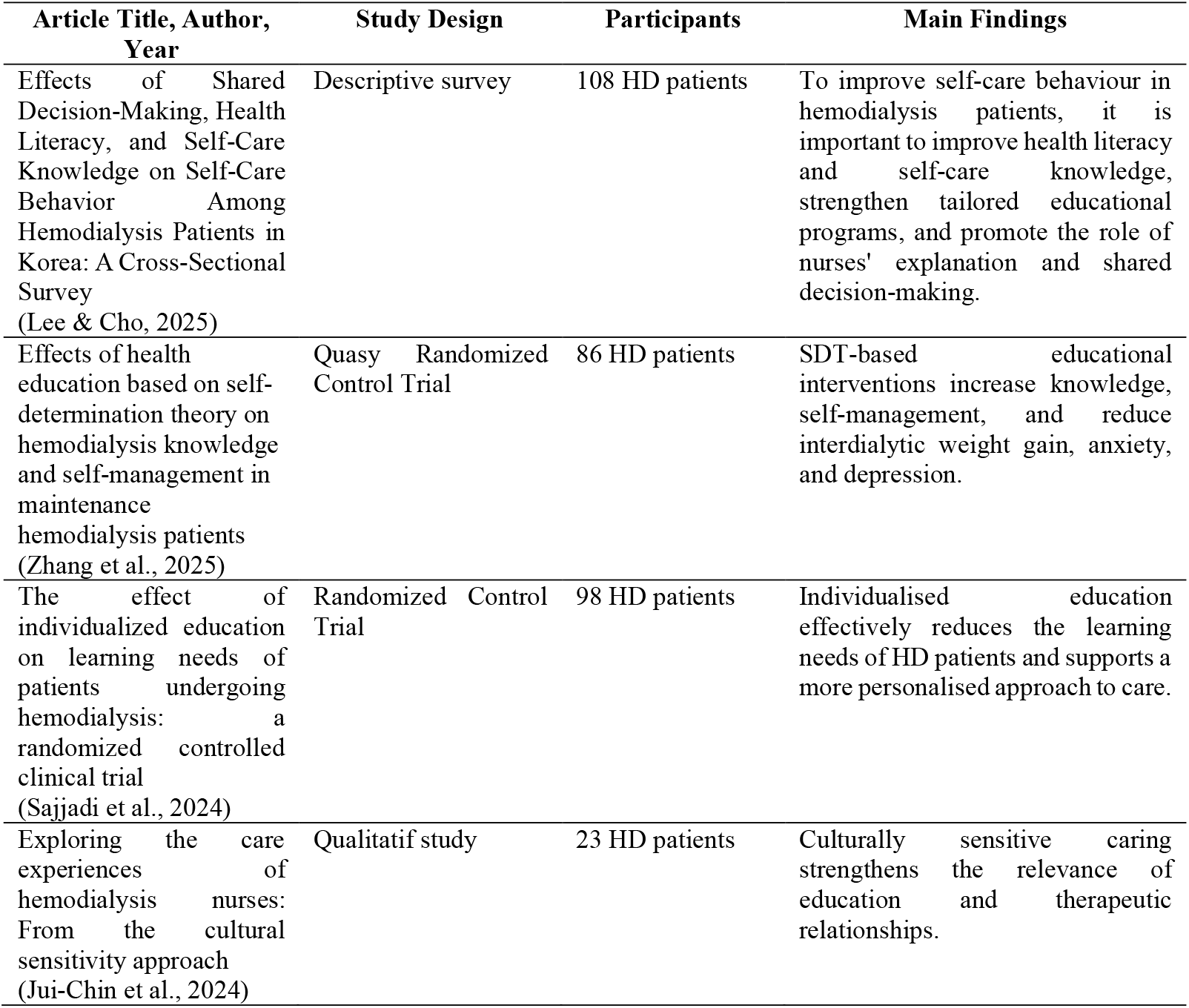
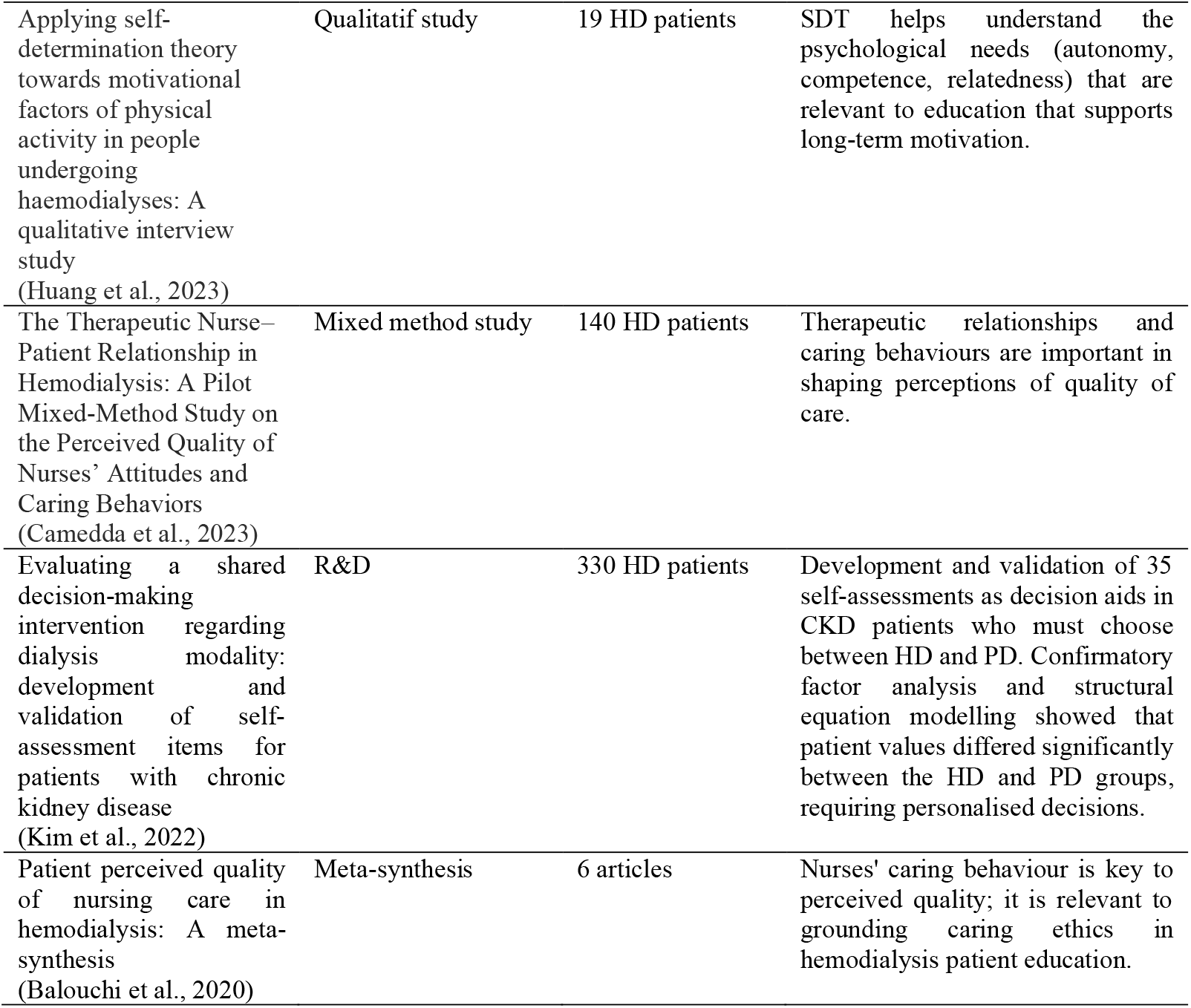
Data Extraction Results

## Discussion

Hemodialysis patient education requires an approach that goes beyond simply conveying medical information; it must position patients as individuals with values, choices, and life experiences. An autonomy perspective is crucial because patients who feel empowered to choose and define their health goals will be more intrinsically motivated to adhere to therapy. Self-Determination Theory explains that supporting basic needs: autonomy, competence, and relatedness, will increase internal motivation (Deci & Ryan, 2000). Research in patients with chronic kidney disease shows that self-efficacy mediates the relationship between knowledge and self-care behaviours, so educational interventions that support autonomy have the potential to improve long-term adherence (Wu et al., 2016).

The theory dominance map diagram (Figure 2) shows that Self-Determination Theory (SDT) occupies the most central position in health education for hemodialysis patients, followed by Caring Ethics and Kantian Autonomy. SDT is powerful because it provides a mechanism for intrinsic patient motivation through fulfilling basic psychological needs: autonomy, competence, and relatedness. However, challenges lie in the cultural context and the healthcare system. In more paternalistic cultures, patient autonomy is often limited, so implementing SDT requires sensitive adaptation to local values and norms.

**Figure 2.**
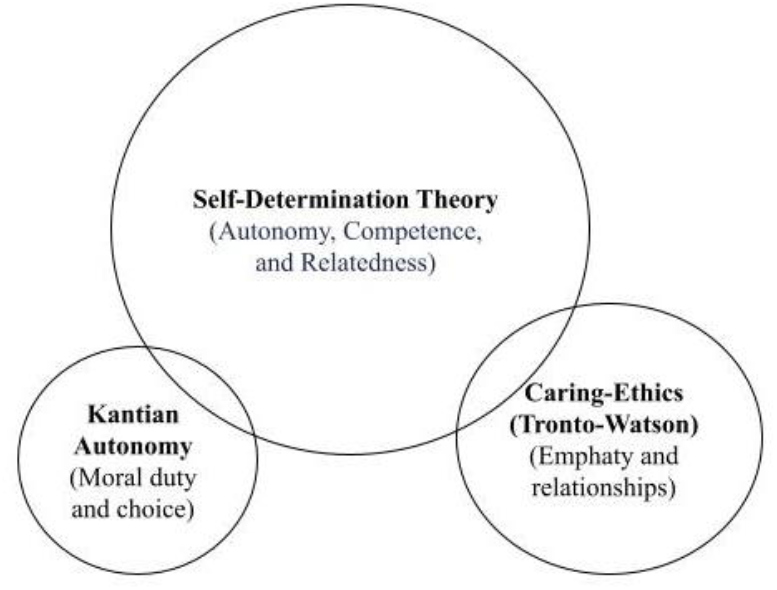
Map of Dominance of Philosophical Theories in Hemodialysis Patient Education

Caring ethics, as outlined by Tronto and Watson, adds strength to the relational and empathetic aspects of education. Recent research emphasises the importance of communication that respects patient values and contextual empowerment (Jui-Chin et al., 2024). The benefit is the creation of a more humane therapeutic relationship, but its limitations include the potential for nurses’ subjective bias and the need for greater time and resources to build deep relationships.

The Kantian principle of autonomy serves as a normative foundation that ensures that health education is not only practical but also ethical. By emphasising the moral obligation to respect patient choice, this theory strengthens professional legitimacy in educational practice. However, its implementation can clash with clinical reality, especially when patient choices align with medical recommendations. Finally, phenomenology provides depth by uncovering subjective patient experiences, often overlooked in technical educational approaches. This perspective enriches practice with a more personal understanding of patients’ suffering, hopes, and meaning in life. A significant challenge is the qualitative and reflective nature of phenomenological methods, making them difficult to practically operationalise within standard educational curricula.

Recent literature analysis reveals four major themes related to the application of philosophy in health education for hemodialysis patients. First, patient autonomy and Shared Decision-Making (SDM) are dominant aspects of health education. Lee & Cho (2025) found that health literacy was the strongest predictor of self-care behaviour, while participation in SDM significantly contributed to patient adherence. This underscores the importance of patient empowerment through education that emphasises choice and autonomy.

These findings underscore the crucial role of philosophy in underpinning health education for hemodialysis patients. The principle of autonomy, grounded in Kantian deontological ethics, emphasises that patients should be viewed as moral agents entitled to make decisions regarding their own care. Human resource practices align with this notion by allowing for active patient participation in choosing disease management strategies.

Second, personalised education has proven effective. Sajjadi et al. (2024) demonstrated through a randomised controlled trial that educational interventions tailored to individual learning needs significantly reduced educational needs, increased understanding, and gave patients a greater sense of control in self-management. Third, caring ethics and cultural sensitivity emerged as important dimensions in nurse-patient interactions. Caring ethics emphasises that education is not merely the transfer of information, but also ethical action that demands care, responsibility, and solidarity.

The philosophy of caring and humanism strengthens the quality of educational interactions. Caring is not simply an act of empathy, but the full presence of healthcare professionals who recognise the dignity of patients in fragile situations. Watson (2008), through his theory of Human Caring, emphasises transpersonal relationships that can create a sense of security and emotional support for patients. Phenomenological studies also confirm that hemodialysis patients perceive care and concern as essential parts of their long-term therapy experience (Shahgholian & Yousefi, 2018). Thus, a caring attitude not only enhances educational acceptance but also helps patients adapt to the challenges of daily life.

Fourth, the quality of nursing relationships is the foundation for effective education. A meta-synthesis by Balouchi et al. (2021) emphasised that patients’ experiences of the quality of nursing care are significantly influenced by nurses’ caring behaviours, which align with Tronto’s caring ethics.

A phenomenological perspective is crucial for exploring the subjective meanings experienced by patients, such as feelings of loss of control, stigma, and anxiety due to physical and social limitations. Qualitative findings indicate that patient experiences should inform the development of educational materials to make them more relevant and meaningful (Shahgholian & Yousefi, 2018). The integration of phenomenological research findings ensures that educational content extends beyond “what patients need to know” to “what matters to patients” within the context of their lives.

The integration of these four perspectives is realised in the form of person-centred hemodialysis patient education, as seen in Figure 3. This educational process extends beyond the transfer of medical information to building relationships, listening to patients’ experiences, and empowering them as key actors in their self-care. Research shows that educational interventions that integrate this philosophical dimension can improve patient adherence to diet, fluid restrictions, medication consumption, and adherence to dialysis sessions (Matteson & Russell, 2010; Murali et al., 2019). Ultimately, this increased compliance leads to an improvement in the patient’s quality of life, both physically and psychosocially, so that education can be seen as a bridge between nursing philosophy and real clinical outcomes.

**Figure 3.**
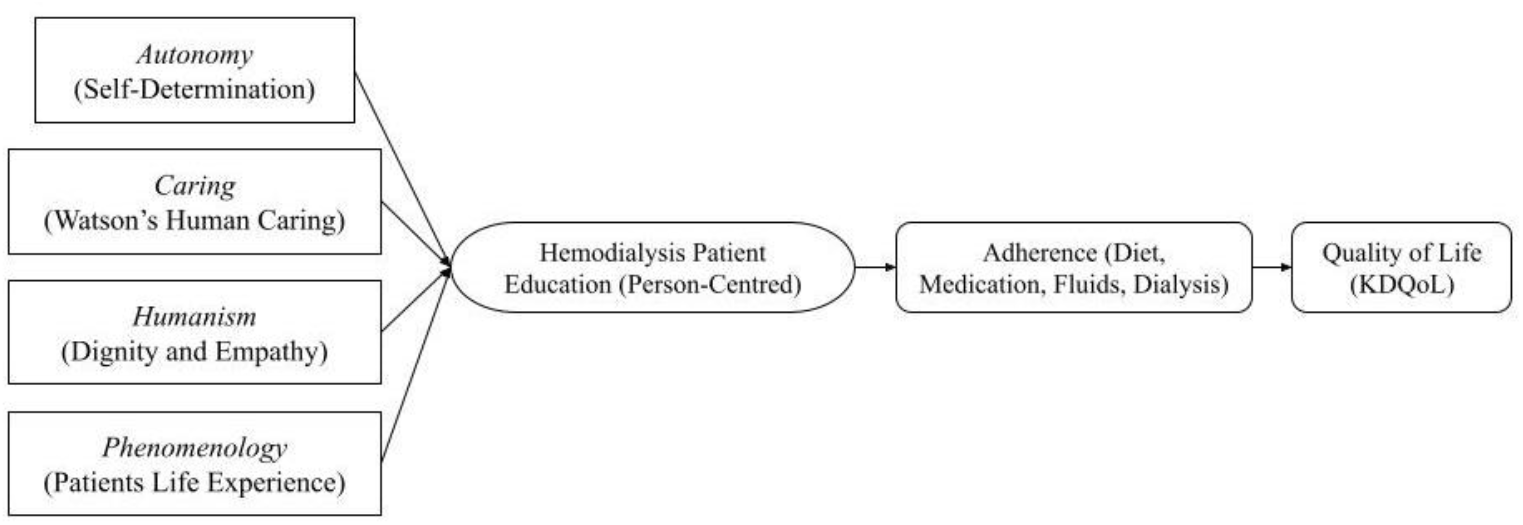
Integration of Philosophical Perspectives in Hemodialysis Patient Education

## Conclusion

This study confirms that hemodialysis patient education cannot be separated from a strong philosophical foundation. Four main perspectives—Self-Determination Theory (SDT), caring ethics, Kantian autonomy, and phenomenology—complement each other in building a more humanistic and ethical educational framework. SDT emphasises the importance of intrinsic motivation through autonomy, competence, and connectedness; caring ethics fosters empathy and relational responsibility; Kantian autonomy emphasises the moral obligation to respect patient dignity; while phenomenology enriches with a deeper understanding of patients’ lived experiences. The integration of these four perspectives encourages educational practices that are not merely technical but also imbued with meaning, ethics, and respect for patients’ subjective experiences.

The implications for nursing practice are the need to develop educational models that emphasise patient autonomy, empathy within the therapeutic relationship, moral respect for individual choices, and an understanding of patients’ personal experiences. Further research is recommended to explore how the integration of these four philosophical perspectives can be operationalised in health education programs and to examine their impact on the independence, quality of life, and adherence of hemodialysis patients across various cultural contexts and healthcare systems.

## Data Availability

All data produced in the present study are available upon reasonable request to the authors

